# The Outcome of COVID-19 Patients with Acute Myocardial Infarction

**DOI:** 10.1101/2020.07.21.20156349

**Authors:** Hassan Al Tamimi, Yasser Alhamad, Fadi Khazaal, Mowahib ElHassan, Hajar AlBinali, Abdul rahman Arabi, Awad Al-Qahtani, Nidal Asaad, Mohammed Al-Hijji, Tahir Hamid, Ihsan Rafie, Ali Omrani, Saad Al Kaabi, Abdullatif Alkhal, Muna Al Maslamani, Mohamed Ali, Murad Alkhani, Mariam AlNesf, Salem Abu Jalala, Salaheddine Arafa, Reem ElSousy, Omar Al Tamimi, Ezeldine Soaly, Charbel Abi Khalil, Jassim Al Suwaidi

## Abstract

**Objectives:** Coronavirus Disease 2019 (COVID-19) is a rapidly expanding global pandemic resulting in significant morbidity and mortality. COVID-19 patients may present with acute myocardial infarction (AMI). The aim of this study is to conduct detailed analysis on patients with AMI and COVID-19.

**Methods:** We included all patients admitted with AMI and actively known or found to be COVID-19 positive by PCR between the 4^th^ February 2020 and the 11^th^ June 2020 in the State of Qatar. Patients were divided into ST-elevation myocardial infarction (STEMI) and Non-STE (NSTEMI).

**Results:** There were 68 patients (67 men and 1 woman) admitted between the 4^th^ of February 2020 and the 11^th^ of June 2020 with AMI and COVID-19. The mean age was 49.1±9 years, 46 patients had STEMI and 22 had NSTEMI. 38% had diabetes mellitus, 31% had hypertension, 16% were smokers, 13% had dyslipidemia, and 14.7% had prior cardiovascular disease. Chest pain and dyspnea were the presenting symptoms in 90% and 12% of patients, respectively. Fever (15%) and cough (15%) were the most common COVID-19 symptoms, while the majority had no viral symptoms. Thirty-nine (33 STEMI and 6 NSTEMI) patients underwent coronary angiography, 38 of them had significant coronary disease. In-hospital MACE was low; 1 patient developed stroke and 2 died.

**Conclusion:** Contrary to previous small reports, in-hospital adverse events were low in this largest cohort of COVID-19 patients presenting with AMI. We hypothesize patient’s demographics and profile including younger age contributed to these findings. Further studies are required to confirm this observation.

**Key questions:** *What is already known on this subject?:*

- COVID-19 patients may present with acute myocardial infarction (AMI).

*What might this study add?:*

- Contrary to previous small reports, most COVID-19 patients presenting with AMI have significant obstructive coronary artery disease and favorable in-hospital outcome.

*How might this impact on clinical practice?:*

- COVID-19 patients presenting with AMI should be treated according to the standard practice.

## Introduction

Beginning December 2019, a sudden outbreak of SARS-CoV-2 (COVID-19) epidemic started in China and spread to many countries around the world. The WHO declared a public health emergency of international concern on the 30^th^ Jan 2020, and to date COVID-19 has become a global pandemic infecting >12 million individuals. The pandemic has led to more than 548,000 fatalities so far (1). COVID-19 mainly affects the respiratory tract, and its clinical manifestations are mostly fever, dry cough, fatigue, and dyspnea. In some cases, the virus can develop into severe pneumonia, acute respiratory distress syndrome (ARDS) and multiple organ dysfunctions (2). Substantial minorities of patients hospitalized with COVID-19 may develop cardiovascular complications.

Acute COVID-19 myocardial injury may occur secondary to acute myocardial infarction (AMI) or myocarditis and can lead to cardiomyopathy, ventricular arrhythmias, hemodynamic instability, and death (3). Furthermore, acute myocardial injury as assessed by troponin release alone appears to be prevalent and is independently associated with worse clinical outcomes among hospitalized COVID-19 patients (2,4-9). The mechanism of acute myocardial injury in COVID-19 patients is unresolved. Several cases of myocarditis have been reported (10-13), while myocardial infarction due to acute plaque rupture in the coronary artery or increased myocardial demand in acute infection phase is the other proposed mechanism (14). AMI among COVID-19 patients may result from severe increase in myocardial demand triggered by infection (type II myocardial infarction) or coagulopathy (15,16). Direct myocardial injury may also occur as a result of alteration of angiotensin converting enzyme 2 (ACE2) signaling pathways as it binds to ACE2 receptors of the myocardium and the lung after entry into the human body (17). Finally, circulating cytokines release during a systemic inflammatory stress may also lead to atherosclerotic plaque rupture (14).

Detailed data about AMI among COVID-19 patients are limited to three preliminary studies (18-20) that included small number of patients and case reports (21-28). They provide incomplete and conflicting findings some of which suggesting high prevalence of non-obstructive coronary artery disease and dismal in-hospital outcome. We aim to evaluate the clinical presentation, demographics, risk factors, angiographic findings and clinical outcomes of AMI in COVID-19 patients in Qatar and review the published literature.

## Methods

### Materials and methods

### Study setting

Qatar is a small country with a population of around 2.6 million: 313,000 Qatari citizens and 2.3 million expatriates. Qatar is classified by the UN as a country of very high human development and is widely regarded as the most advanced Arab state for human development (29,30). The study was conducted at Hamad Medical Corporation, Doha, Qatar which includes a network of hospitals: Hamad General Hospital, Heart Hospital, Wakra Hospital, Alkhor Hospital, Cuban Hospital (COVID-19 hospital) and Hazm Mebaireek Hospital (COVID-19 hospital). The Heart Hospital is the only tertiary referral cardiac center in the country, with a total of 120 beds. All invasive cardiac procedures (cardiac catheterization procedures and cardiac surgeries) are performed at the Heart Hospital (31); prior to the COVID-19 pandemic all STEMI patients were treated with primary percutaneous coronary intervention (PPCI), however during the pandemic occasionally STEMI-patients were treated with fibrinolytic therapy instead of PPCI according to the discretion of the treating physician. During the pandemic, hemodynamically stable and chest pain free COVID-19 patients with Non-ST elevation myocardial infarction (NSTEMI) were managed with conservative medical therapy and planned outpatient coronary angiography in order to reduce the risk of transmission of disease to hospital staff and other hospitalized cardiac patients. Once COVID-19 patients with AMI got treated and stabilized at the Heart Hospital subsequently they were transferred to COVID hospital facilities for further therapy.

The first case of COVID-19 in Qatar was confirmed on the 27^th^ February 2020. As of the 3^rd^ June 2020, Qatar has the highest number of confirmed cases per capita in the world, and the 2nd highest total of confirmed cases in the Arab world after Saudi Arabia at 220,144. The total diagnosed patients were 106,308 with total deaths reported at 154 (0.144% fatality rate). The total number of tests performed in Qatar stands at 438,990 (32).

Retrospective detailed analysis of all hospitalized patients with AMI and COVID-19 in Qatar between February 27 (date of first COVID-19 case in Qatar) and June 12, 2020 was made. All patients hospitalized at the Heart Hospital routinely get COVID-19 PCR testing. Ethical clearance was taken from MRC Research Committee, HMC for the analysis and publication of the study. ClinicalTrials.gov Identifier: NCT04430374.

## Definition

COVID-19 was confirmed by reverse-transcription polymerase-chain reaction (PCR) assay. Patients with COVID-19 had a confirmed diagnosis based on the identification of SARS-CoV-2 on nasal/pharyngeal swab. All of these patients were managed as COVID-19 positive as per center policy. Diagnosis of the different types of AMI and definitions of data variables were based on the American College of Cardiology (ACC) clinical data standards (8). Use of adjunct therapy during hospitalization was recorded for every patient. The presence of diabetes mellitus (DM) was determined by the documentation of DM diagnosis that had been treated with medications or insulin in the patient’s medical record. The presence of dyslipidemia was determined by the demonstration of a fasting cholesterol > 5.2 mmol/L or any history of dyslipidemia treatment in the patient’s medical record. Chronic renal impairment was defined as creatinine >1.5 upper normal range. The presence of hypertension (HTN) was established if there were any documentation of HTN in the medical record or if the patient was taking anti-hypertensive treatment. Smoking history was defined as those who are currently smoking and previous cardiovascular disease was defined with any previous cardiovascular disorders (31).

Primary outcome was in-hospital mortality. Secondary outcomes were major adverse cardiovascular events (MACE), including, recurrent myocardial infarction, stroke, decompensated heart failure, sustained ventricular arrhythmia, atrioventricular heart block, and hemodialysis initiation for acute renal failure. In-hospital outcomes were reported for the entire stay at the Heart Hospital and other COVID-19 facility hospitals in Qatar.

### Statistical Analysis

Data were presented in the form of frequency and percentages for categorical variables and mean ± standard deviation (SD) or Standard Error (SE). Due to the small sample size, only descriptive analysis was performed.

## Results

### Clinical Presentation and Baseline patient characteristics (Tables 1&2)

Between the 4^th^ February 2020 and the 11^th^ June 2020, 68 patients (67 men and 1 woman) were admitted with COVID-19 and AMI. The mean age of patients was 49.1±9 years, 65 patients were South Asians, 2 Arabs and one was African. Forty-six patients had STEMI (28 anterior and 18 inferior) and 22 patients had NSTEMI (Table 1).

**Table 1.** Baseline Clinical Characteristics.

| No |  | All (%) | STEMI (%) | NSTEMI (%) |
| --- | --- | --- | --- | --- |
| Total Number |  | 68 | 46 (67.6%) | 22 (35.4%) |
| Gender | Male | 67 (98.5%) | 46 (100%) | 21 (95%) |
|  | Female | 1 (1.5%) | 0 | 1 (5%) |
| Age (years) |  | 49+/-9 | 49.5+/-8.8 | 48+/-8 |
| Ethnicity |  | Arab =2 (3%)<br>Asian= 65(95%)<br>Other= 1(1.5 %) | Arab =1(2 %)<br>Asian= 44(96%)<br>Other=1(2%) | Arab =1(4.5 %)<br>Asian= 21(95%)<br>Other=0 (1.5%) |
| Cardiovascular risk profile | Previous Cardiovascular Disease | 10 (14.7%) | 6(8.7%) | 4(18%) |
|  | Diabetes Mellitus | 26(38%) | 18(39%) | 8(36%) |
|  | Hypertension | 21(31%) | 16(35%) | 5(23%) |
|  | Smoking | 11(16%) | 10(22%) | 1(4.5%) |
|  | Dyslipidemia | 9(13%) | 6(13%) | 3(13%) |
|  | Chronic Kidney Disease | 0 | 0 | 0 |
| Cardiac Symptoms | Chest pain | 61(90%) | 40(87%) | 21(95%) |
|  | Dyspnea | 8(12%) | 7(15%) | 1(5%) |
|  | Palpitation | 1(1.5%) | 1(2%) | 0 |
|  | Syncope | 3(4%) | 3(7%) | 0 |
|  | Out of Hospital Cardiac Arrest | 0(%) | 0 | 0 |
| Blood Pressure | SBP | 133±25 | 129±26 | 140±20 |
|  | DBP | 83±14 | 81±15 | 86±10 |
| Respiratory rate |  | 20±7 | 21±7 | 19±3.4 |
| Body mass index |  | 25±4 | 25±4 | 26±3.5 |
| Shared Accommodation (Either with family or worker housing) |  | 56(82%) | 37(80%) | 19(86%) |
| COVID Symptoms | Asymptomatic | 53(78%) | 34(74%) | 19(86%) |
|  | Fever | 10(15%) | 9(20%) | 1(5%) |
|  | Cough | 11(16%) | 10(22%) | 1(5%) |
|  | Sore throat | 3(4%) | 3(6.5%) | 0(0%) |
|  | Myalgia | 4(6%) | 3(6.5%) | 1(5%) |
|  | Shortness of Breath | 6(9%) | 6(13%) | 0 |
|  | Loss of taste or smell | 1(%) | 1(2%) | 0 |
|  | Diarrhea | 0 | 0 | 0 |

STEMI=ST-elevation myocardial infarction, NSTEMI=Non-ST elevation myocardial infarction, CAD=cardiovascular disease, SBP=systolic blood pressure, DBP=diastolic blood pressure.

Out of the total 68 patients; 4 patients were known to be COVID-19 positive and were under treatment in a COVID-19 facility when they developed STEMI; the remaining patients were diagnosed with COVID-19 at the time of presentation with AMI. Risk factors analysis (Table 1) showed 14.7% had prior cardiovascular disease, 38% of patients had diabetes mellitus, 31% had hypertension, 16% were smokers, and 13% had dyslipidemia. Chest pain and dyspnea were the presenting symptoms in 90% and 12% of patients, respectively. Fever (15%) and cough (16%) were the most prevalent COVID-19 symptoms, while the majority of patients had no viral symptoms. Most patients had normal white blood cell count and no evidence of lymphopenia on admission (Table 2).

**Table 2.** Laboratory Investigations.

|  | All |  |  | STEMI |  | NSTEMI |  |
| --- | --- | --- | --- | --- | --- | --- | --- |
|  | Normal Range | Mean | SE | Mean | SE | Mean | SE |
| WBC at admission, | (4-10)<br>X10<sup>3</sup>/uL | 9.9 | 0.48 | 10.63 | 0.67 | 8.41 | 0.37 |
| Lymphocytes at admission, | (1-3)<br>X10<sup>3</sup>/uL | 1.9 | 0.11 | 1.76 | 0.14 | 2.22 | 0.18 |
| HB at admission, | (13-17)<br>gm/dL | 14.23 | 0.19 | 14.17 | 0.23 | 14.35 | 0.3 |
| Platelet at admission, | (150-400)<br>X10<sup>3</sup>/uL | 318.8 | 16.4 | 330.4 | 22 | 301 | 23.5 |
| HB at Discharge, | (13-17)<br>gm/dL | 15.21 | 1.82 | 15.71 | 2.59 | 14.02 | 0.39 |
| Glucose (Random), | (Fasting<br>3.3-5.1)<br>mmol/L | 10.62 | 0.89 | 10.9 | 1.17 | 10.05 | 1.31 |
| HbA1C, % | 4.0-5.5% | 7.5 | 0.29 | 7.52 | 0.35 | 7.47 | 0.54 |
| Creatinine at Admission, | 62-106<br>umol/L | 86.96 | 3.79 | 87.84 | 4.98 | 85.14 | 5.57 |
| Creatinine At Discharge, | 62-106<br>umol/L | 107.88 | 17.04 | 115.44 | 24.37 | 90.53 | 4.85 |
| Cholesterol, | Less than<br>5.18<br>mmol/L | 4.31 | 0.15 | 4.1 | 0.18 | 4.78 | 0.25 |
| Triglycerides, | Less than<br>3.88<br>mmol/L | 1.67 | 0.12 | 1.62 | 0.14 | 1.78 | 0.22 |
| HDL Cholesterol | More than<br>1.55<br>mmol/L | 0.82 | 0.02 | 0.8 | 0.03 | 0.86 | 0.04 |
| LDL Cholesterol | Less than<br>2.59<br>mmol/L | 2.7 | 0.14 | 2.5 | 0.17 | 3.11 | 0.23 |
| HS Troponin T | 3.0-15<br>ng/L | 4424.49 | 643.11 | 5942.84 | 865.03 | 1318.77 | 260.52 |
| Creatine Kinase | 50-200<br>U/L | 773.12 | 179.79 | 1016.73 | 254.49 | 367.11 | 161.1 |
| ProBNP, U/L | Below 125<br>pg/mL | 1223.38 | 314.02 | 1442.88 | 447.56 | 770.67 | 254.32 |
| LDH | 125-225U/L | 551.91 | 66.21 | 698.54 | 92.27 | 295.31 | 29.15 |
| D dimer, mg/L | 0-0.49<br>mg/L | 4.42 | 1.84 | 2.49 | 0.91 | 9 | 5.84 |
| Fibrinogen | 2.0-4.1<br>gm/L | 4.9 | 0.25 | 5.2 | 0.3 | 4.23 | 0.37 |
| CRP, mg/L | 0-5 mg/L | 54.76 | 8.93 | 65.51 | 11.72 | 33.26 | 12.06 |
| Ferritin, ug/L | 24-336<br>mcg/L | 1223.61 | 652.07 | 1605.4 | 944.19 | 379.67 | 63.5 |
| Interleukin 6 | ≤7 pg/mL | 730 | 711.69 | 1271.5 | 1242.85 | 8 | 1.53 |
| Procalcitonin | <0.5<br>ng/mL<br>low risk<br>for sepsis<br>>2.0<br>ng/mL<br>high risk<br>for sepsis | 5.53 | 4.54 | 7.37 | 6.23 | 0.63 | 0.37 |
| Lactic acid | 0.5-2.2<br>mmol/l | 2.27 | 0.56 | 2.43 | 0.76 | 1.92 | 0.59 |
| INR |  | 1.12 | 0.03 | 1.14 | 0.04 | 1.07 | 0.02 |
| Potassium | 3.5-5.1<br>mmol/L | 4.15 | 0.07 | 4.09 | 0.1 | 4.27 | 0.08 |
| Sodium | 136-145<br>mmol/L | 136.99 | 0.41 | 137.09 | 0.54 | 136.77 | 0.63 |
| Magnesium | 0.6-0.9<br>mmol/L | 0.83 | 0.01 | 0.82 | 0.02 | 0.85 | 0.02 |

SE=standard error, WBC=white blood cell count, HB=hemoglobin, HDL=high density lipoprotein, LDL=low density lipoprotein, CRP=C Reactive Protein, INR=International Normalization Ratio.

### Management and Outcome (Table 3-5)

Most patients received evidence-base therapy. Of the 46 STEMI patients; 38 (82.6%) patients underwent reperfusion therapy (32 PPCI and 7 fibrinolytic therapy), while 8 patients did not undergo reperfusion therapy due to late presentation. One patient underwent successful rescue PCI after failed reperfusion by fibrinolytic therapy. Most of the NSTEMI patients (n=16) were managed initially conservatively with medical therapy alone, while 6 patients underwent coronary angiogram due to hemodynamic instability or persistent chest pain. Those 6 patients underwent subsequent revascularization by PCI (n=3) or CABG (n=3). In total, 39 (57.3%) AMI patients (STEMI & NSTEMI) underwent coronary angiography showing obstructive coronary artery disease (CAD) with at least one culprit lesion in all with the exception of one STEMI patient who had mild non-obstructive CAD (Table 3).

**Table 3.** Coronary Diagnostic and therapeutic intervention.

|  |  | All | STEMI | NSTEMI |
| --- | --- | --- | --- | --- |
| Coronary angiogram |  | N=39 | N=33 | N=6 |
| Culprit lesion | LMCA | 5 (7%) | 3(7%) | 2(9%) |
|  | LAD | 17(25%) | 13(28%) | 4(18%) |
|  | D (any D) | 4(6%) | 3(7%) | 1(5%) |
|  | LCx | 7(10%) | 4(9%) | 3(14%) |
|  | OM (any OM) | 12(18%) | 9(20%) | 3(14%) |
|  | RCA | 8(12%) | 7(15%) | 1(5%) |
|  | PDA | 0 | 0 | 0 |
|  | Stent thrombosis at presentation | 3(4%) | 2(4%) | 1(5%) |
|  | Stent thrombosis as complication | 1(1%) | 1(2%) | 0 |
| No significant CAD by coronary angiography |  | 1/39 (2.6%) | 1/33(3.0%) | 0 |
| 1VD by coronary angiography |  | 17/39 (43.6%) | 15/33 (45.5%) | 2/6 (33.3) |
| 2VD by coronary angiography |  | 8/39 (20.5%) | 7/33 (21.2%) | 1/6 (16.7%) |
| 3VD by coronary angiography |  | 12/39 (30.8%) | 9/33 (27.3%) | 3/6 (50%) |
| Late Presentation |  | 8 (12%) | 8 (17%) | NA |
| Fibrinolytic therapy |  | 7 (10%) | 7 (15%) | NA |
| Total PCI |  | 35 (51.5%) | 32 (69.6%) | 3 (5%) |
| CABG |  | 3(4.4%) | 0 | 3(14%) |
| Mechanical Circulatory support |  | 0 | 0 | 0 |
| Transvenous Temporary Pacing |  | 1(1.5%) | 1(2%) | 0 |
| Echocardiography | Number of patients | 34 (50%) | 19 (41.3%) | 15 (68.2%) |
|  | EF | 46±14 | 43.6±11 | 51±12 |

LMCA=Left main coronary artery disease, LAD=left anterior descending artery, RCA=right coronary artery, LCx=left circumflex artery, D=Diagonal artery, EF=ejection fraction, CABG=Coronary artery bypass grafting, OM=obtuse marginal artery,

Overall in-hospital MACE were low. In-hospital adverse events were reported in 9 patients only (Table 4); 4 patients developed decompensated heart failure, 1 had cardiogenic shock, 1 patient had stroke, 1 patient had stent thrombosis and 2 patients died. Of the two mortalities, one was a 57-year old Asian man with diabetes and hypertension who presented with late inferior myocardial infarction, cough, and fever. The patient was managed conservatively including therapeutic doses of low molecular weight heparin and dual antiplatelet therapy. Three days later he deteriorated developing cardiogenic shock and acute renal failure. The patient expired on day 7 of hospitalization. The second patient was also 57-year old Asian man who presented with inferior STEMI, complete heart block and severe COVID-19 pneumonia. Temporary pacemaker was placed, and coronary angiography was performed showing severe 3-vessel disease including significant left main coronary artery disease. The patient underwent balloon angioplasty of the culprit artery (RCA). Cardiovascular surgery deemed the patient not candidate for surgery and unfortunately, he progressively developed multi-organ failure and expired on hospitalization day 14. The patient who developed stroke was 44-year old man who underwent PPCI for anterior STEMI and COVID-19 pneumonia. Coronary angiography at that time demonstrated extensive thrombus in the LAD. His procedure was complicated by migration of thrombus into the left circumflex artery after ballooning. The patient was further managed with aspiration thrombectomy, and intravenous infusion of heparin and glycoprotein IIb/IIIa inhibitors. His post-intervention LVEF was 32% without evidence of LV thrombus. The patient stabilized and was transferred to COVID-19 facility to complete his therapy, however on day 8 after STEMI he developed right leg weakness. CT Head and CT Cerebral angiogram were performed without evidence of hemorrhagic or ischemic defects. The patient subsequently recovered completely and discharged home.

**Table 4.** In-hospital and discharge medications.

| Medications |  |  |  |
| --- | --- | --- | --- |
| Aspirin | In hospital =68 (100%)<br>At discharge =54(79%) | In hospital =46(100%)<br>At discharge =37(80%) | In hospital =22(100%)<br>At discharge =17(77%) |
| Clopidogrel | In hospital =64(94%)<br>At discharge =50(74%) | In hospital = 43(93%)<br>At discharge = 34(74%) | In hospital = 21(95%)<br>At discharge = 16(73%) |
| Ticagrelor | In hospital =1(1.5%)<br>At discharge =2(3%) | In hospital = 1(2%)<br>At discharge =2(4%) | In hospital =0<br>At discharge =0 |
| Beta blockers | In hospital =55(81%)<br>At discharge =49(72%) | In hospital = 37(80%)<br>At discharge = 34(74%) | In hospital = 18(82%)<br>At discharge = 15(68%) |
| ACE/ARB | In hospital =31(46%)<br>At discharge =27(40%) | In hospital = 23(50%)<br>At discharge =20(43%) | In hospital = 8(36%)<br>At discharge =7(32%) |
| Statin | In hospital =68(100%)<br>At discharge =52(76%) | In hospital = 46(100%)<br>At discharge = 36(78%) | In hospital = 22(100%)<br>At discharge =16(73%) |
| Nitrate | In hospital =34(50%)<br>At discharge =26(38%) | In hospital =21(46%)<br>At discharge =16(35%) | In hospital =13(59%)<br>At discharge =10(45%) |
| COVID Medications |  |  |  |
| Hydroxychloroquine | 35(51%) | 21(46%) | 14(64%) |
| Azithromycin | 48(71%) | 33(72%) | 15(68%) |
| Oseltamivir | 16(24%) | 13(28%) | 3(14%) |
| Tocilizumab | 2(3%) | 2(4%) | 0 |
| Lopinivir/Ritonavir | 4(6%) | 2(4%) | 2(9%) |
| Draunavir/Cobicisat | 0 | 0 | 0 |

ACE=angiotensin converting enzyme inhibitors, ARB=angiotensin receptor blocker.

**Table 5.** In-hospital Outcome.

| No |  | All | STEMI | NSTEMI |
| --- | --- | --- | --- | --- |
| In-hospital Outcome |  |  |  |  |
| Acute Heart Failure |  | 4 (6%) | 3(6%) | 1 (%) |
| Cardiogenic shock |  | 1(1.5%) | 1(2%) | 0 |
| Arrythmia | Atrial Tachyarrhythmia | 0 | 0 | 0 |
|  | Ventricular tachyarrhythmia | 6(8.8%) | 6(13%) | 0 |
| Atrioventricular block |  | 7(10%) | 7(15%) | 0 |
| AV Block requires Temporary Pacing |  | 2(3%) | 2(4%) | 0 |
| Re infarction |  | 2(3%) | 2(4%) | 0 |
| Acute renal failure requires hemodialysis |  | 3(4.4%) | 3(6%) | 0 |
| Major Bleeding |  | 1(1.5%) | 1(2%) | 0 |
| Left Ventricular thrombus |  | 0 | 0 | 0 |
| Thrombotic events /Stroke |  | 1(1.5%) | 1(2%) | 0 |
| Invasive Mechanical Ventilation |  | 4(6%) | 4(9%) | 0 |
| All-cause mortality |  | 2(3%) | 2(4%) | 0 |

### AMI in Known COVID-19 patients under treatment

Four of the 68 patients in the study developed STEMI (all anterior) with symptoms while been treated in a COVID-19 facility for a variable duration of time 8-11 days (ages; 28, 43, 54 and 55 years old). All of them were subsequently transferred to the Heart Hospital and underwent coronary angiography with successful PCI to the LAD. Interestingly the youngest patient (28-year old patient) had highly elevated platelet count 914,000 on the day of presentation and gradually resolved to baseline. The patient had no other parameters to suggest inherited coagulopathy. All four patients were discharged home after successful revascularization.

### Stent Thrombosis, AMI and COVID-19

Three COVID-19 patients presented with AMI due to stent thrombosis; Two patients presented with NSTEMI and were found to have very-late stent thrombosis, COVID-19 was diagnosed at the time of presentation; a 45-year old Asian man who presented with obtuse marginal coronary artery stent thrombosis, the other patient was a 58-year old Asian man who presented with thrombosis of LAD stent placed 4 years earlier. The third patient with stent thrombosis initially presented with inferior STEMI and underwent PPCI of the RCA, after stabilization he transferred to a COVID facility, unfortunately antiplatelet therapy was prematurely discontinued and he was transferred back to the Heart Hospital with stent thrombosis 14 days later requiring repeat intervention. All three patients were discharged home after completion of the treatment.

## Discussion

The current study provides detailed analysis of the ***largest*** cohort of patients with COVID-19 presenting with AMI to-date. Our patients were younger with lower cardiovascular risk profile when compared to previously published small reports (18-20). In contrast to most of previous reports (18,19) which suggested high prevalence of non-obstructive coronary artery disease among AMI COVID-19 patients and worse outcome, significant obstructive coronary angiography was highly prevalent in our cohort, moreover, in-hospital adverse events were very low with only 1 stroke and 2 deaths reported.

The COVID-19 pandemic had affected cardiovascular diseases and their management in multiple ways. First, the escalation of this health crisis has led to cancellation of elective cardiovascular procedures and outpatient visits, due to the concern of disease transmission among healthcare providers and other patients, as well as to optimize resource allocation. Second, several investigators reported significant decline in AMI rates during the pandemic. De Rose et al (34) reported significant reduction in AMI admissions during the pandemic across Italy, with a parallel increase in fatality and complication rates. Similar observations were reported from China, UK and USA (35). Bainey suggested this drop in AMI rates perhaps partly due to the patients unwilling to access the emergency medical system or risk hospital exposure to COVID-19 (36). Third, several experts suggested amidst the pandemic that the STEMI networks needed to re-evaluate their STEMI care systems to best balance the needs of the patients and the health and safety of their healthcare providers. In China, fibrinolytic therapy was suggested as the primary reperfusion therapy, while in the United States, several experts had differing opinions in regard to fibrinolytic therapy use versus PPCI (36-38). Daniels et al. (38) suggested in those challenging and unprecedented times fibrinolytic therapy as primary reperfusion therapy might be a reasonable option.

### Clinical Characteristics of COVID-19 AMI Patients and Outcome

To date there are only three studies one from Lombardy (Italy) (18), the other from New York city (19) and more recently from the UK (20) that provided detailed information of AMI among COVID-19 patients, in addition to a number of case reports from around the world (Table 6). Stefanini et al in a retrospective analysis reported 29 patients who underwent coronary angiography for STEMI in Lombardy, Italy between February 2020 and March 2020, most had unknown COVID status at the time of presentation. The mean age of patients was 68±11 years, 8 patients (28.6%) were women and 3 (10.7%) had a prior myocardial infarction. The majority presented with typical chest pain with or without dyspnea (78.6%), 6 patients (21.4%) had dyspnea without chest pain (ours was 11%). In contrast to ours, all Lombardy patients underwent urgent coronary angiography, and none were treated with fibrinolysis. 39.3% did not have obstructive coronary artery disease. The mortality rate of their cohort was high 11 (39.3%) patients died.

**Table 6.** Reported studies of acute coronary syndrome COVID-19 patients.

| No | Authors | Country | Total no of patients | Age (Yrs) | Men (%) | DM (%) | HTN (%) | CKD (%) | Prior CV (%) | STEMI | NSTEMI | PPCI | Fibrinolytic | No significant CAD (%) | Others | Mortality (%) |
| --- | --- | --- | --- | --- | --- | --- | --- | --- | --- | --- | --- | --- | --- | --- | --- | --- |
| 1 | Stefanine (18) | Lombardy (Italy) | 28 | 68+11 | 71.4 | 32.1 | 21.4 | 28.6 |  | All | - | Yes | No | 39.3 |  | 39.3 |
|  | Banglore (19) | New York (USA) | 18 | 65(54-73) | 83 | 35 | 65 | 6 |  | All |  |  | 1/18 | 33% □ | Poor prognosis |  |
| 2 | Xuan (20) | China * | 1 | 70 | Yes | Yes | Yes | No | No | Yes | No | Yes | No | No | Lung transplantation for COVID | No |
| 5 | Lacour T(21) | France | 1 | 68 |  |  |  |  |  | Yes | No | Rescue | Failed | No | 2 episodes of stent thrombosis | Died |
| 6 | Yasushi Ueki (22) | Switzerland | 1 | 83 |  |  |  |  |  | Yes |  | Yes |  |  |  |  |
| 7 | Siddamreddy S (23) |  | 1 | 61 | No | Yes | Yes |  | Yes | Inferior | No | Yes | No | No | Stent thrombosis | No |
| 8 | Zhichao Xiao (24) 3 patients | China | 1 | 62 | Yes | No | No |  | No | Yes | No | No | Yes |  | Successful | Yes |
|  |  | China | 1 | 42 | Yes | Yes | Yes |  | Yes¶ | Yes | No | Yes | No | No | Stent thrombosis | Yes |
|  |  | China | 1 | 78 | Yes |  |  |  |  | Yes | No | Yes | No |  |  | Yes |
| 9 | Hinterseer (25) | Germany | 1 | 65 | Yes | Yes | Yes | No | Yes | * | * | Yes | No | No | Stent thrombosis | Died |
| 10 | Seif (26) | United Kingdom | 1 | 58 | No | ? | ? | ? | ? | Inferior | No | Yes | No | No | Failed because of extensive thrombosis. | Died |
|  | Current study | Qatar | 68 | 49 | 67 (98.5%) | 26(38%) | 21(31%) | 0 | 10 (14.7%) | 46(67.7%) | 22(35.4%) | 31(67.4%) | 7 (15%) | 48% | 4 stent thrombosis 2 deaths | 8 |

DM=diabetes mellitus, HTN=hypertension, CKD=chronic kidney disease, Prior CV=prior cardiovascular disease, PPCI=primary percutaneous coronary interventions, \*RBBB and ST-elevation aVR, ¶=primary PCI LAD 20 days prior to current admission, □ = 9 patients had coronary angiography. CAD=coronary artery disease.

Banglore’s et al preliminary study from 6 New York Hospitals reported 18 confirmed COVID-19 patients presenting with ST-segment elevation on ECG indicating potential AMI. Their median age was 63 years, 88% were men and only 33% of patients had chest pain around the time of ST-segment elevation. A total of 9 patients (50%) underwent coronary angiography; 6 of these patients (67%) had obstructive disease, and 5 (56%) underwent percutaneous coronary intervention (1 after the administration of fibrinolytic agents). Thirteen out of 18 patients died during hospitalization. More recently, an observational study that included 115 consecutive patients who underwent PPCI for STEMI from Barts Heart Center in the UK was reported (20); there were 39 COVID-19 patients and 76 non-COVID-19 patients. The mean age of patients was 62 years and 47% of patients were from black, Asians or ethnic minority groups. COVID-19 patients were more likely to be diabetic and have hypertension. There was evidence of higher thrombogenicity in the COVID-19 group with significantly higher rates of multi-vessel thrombosis and stent thrombosis and in contrast to the other two reports the majority underwent PCI suggesting the presence of significant obstructive CAD. COVID-19 patients had higher incidence of cardiac arrest and a trend of higher in-hospital mortality rate (17.9% vs 6.5% in Non-COVID-19 patients, *p*=0.10). This in-hospital mortality was much lower than previous reports (18,19).

Our patients’ populations are almost 10 years younger than these three reports (18-20), which may explain the improved survival to discharge when compared to their cohorts. Moreover, most of our patients had no COVID-19 symptoms, severe respiratory distress, or other evidence of coagulopathy at the time of presentation. Contrary to both Stefanini (18) and Banglore (19) reports that suggested high prevalence of non-obstructive coronary artery disease (39% and 33%), *only* 1 out of 39 patients referred for angiography had no culprit lesion (s) in our study which is consistent with the recent observations by Choudry et al (20). Twenty-four out of 68 patients did not undergo angiography in our study. These patients were felt to have intermediate-low risk presentations by their treating providers and hence coronary angiography was deferred until resolution of their COVID-19 infection. Recently in order to improve COVID-19 AMI patients’ management a dedicated diagnostic pathway has been outlined in a consensus statement from the Society of Cardiovascular Angiography and Interventions (SCAI), American College of Cardiology (ACC), and the American College of Emergency Physicians (ACEP) (39) which has been adopted by our institution.

### Limitation of the study

Our study is constrained by the limitations inherent in all studies of observational design. We also acknowledge that this is an early report on a relatively small number of patients, however, to the best of our knowledge this is the *largest reported cohort to date*. Another limitation of the study is the lack of standard definition of COVID-19 associated myocardial infarction which has not been defined to date. Fourth, 35% of our AMI patients did not undergo coronary angiography because their treated physicians regarded them as low-risk cases and decided to have coronary angiography performed later once their COVID-19 infection resolved. Therefore, we do not have the angiographic data for this group at present. The current study highlights the importance of developing large-scale registries to accurately describe the association and outcome of COVID-19 AMI patients. Multiple registries are underway including North American COVID-19 ST-segment elevation myocardial infarction registry (NACMI). The registry is organized by the Society for Cardiovascular Angiography and Interventions (SCAI) and The Canadian Association of Interventional Cardiology (CAIC) in conjunction with the American College of Cardiology Interventional Council. (39). Finally, long-term data is not available.

## Conclusion

Contrary to previous small reports, the majority of COVID-19 patients presenting with AMI have significant obstructive coronary artery disease and overall excellent in-hospital outcome. We hypothesize patient’s profile including younger age contributed to these findings. Further studies are required to confirm this observation. Further studies are required to confirm this observation.

## Data Availability

All data are avalaible.

## Abbreviations

COVID-19: coronavirus 2019;
EMS: emergency medical services;
ICU: intensive care unit;
PCI: percutaneous coronary; intervention;
STEMI: ST-elevation myocardial infarction
STEMI: ST-elevation myocardial infarction=Non-ST elevation myocardial infarction
NACMI: North American COVID-19 ST-segment elevation myocardial infarction registry
SCAI: Society for Cardiovascular Angiography and Interventions
CAIC: Canadian Association of Interventional Cardiology
ACEP: American College of Emergency Physicians.

